# Network localization of pediatric lesion-induced dystonia

**DOI:** 10.1101/2024.04.06.24305421

**Authors:** Rose Gelineau-Morel, Nomazulu Dlamini, Joel Bruss, Alexander Li Cohen, Amanda Robertson, Dimitrios Alexopoulos, Christopher D. Smyser, Aaron D. Boes

## Abstract

**Objective:** Dystonia is a movement disorder defined by involuntary muscle contractions leading to abnormal postures or twisting and repetitive movements. Classically dystonia has been thought of as a disorder of the basal ganglia, but newer results in idiopathic dystonia and lesion-induced dystonia in adults point to broader motor network dysfunction spanning the basal ganglia, cerebellum, premotor cortex, sensorimotor, and frontoparietal regions. It is unclear whether a similar network is shared between different etiologies of pediatric lesion-induced dystonia.

**Methods:** Three cohorts of pediatric patients with lesion-induced dystonia were identified. The lesion etiologies included hypoxia, kernicterus, and stroke versus comparison subjects with acquired lesions not associated with dystonia. Multivariate lesion-symptom mapping and lesion network mapping were used to evaluate the anatomy and networks associated with dystonia.

**Results:** Multivariate lesion-symptom mapping showed that lesions of the putamen (stroke: r = 0.50, p <0.01; hypoxia, r = 0.64, p <0.001) and globus pallidus (kernicterus, r = 0.61, p <0.01) were associated with dystonia. Lesion network mapping using normative connectome data from healthy children demonstrated that these regional findings occurred within a common brain-wide network that involves the basal ganglia, anterior and medial cerebellum, and cortical regions that overlap the cingulo-opercular and somato-cognitive-action networks.

**Interpretation:** We interpret these findings as novel evidence for a unified dystonia brain network that involves the somato-cognitive-action network, which is involved in higher order coordination of movement. Elucidation of this network gives insight into the functional origins of dystonia and provides novel targets to investigate for therapeutic intervention.

## Introduction

Dystonia is a movement disorder characterized by involuntary intermittent or sustained muscle contractions leading to twisting or repetitive movements and abnormal postures.^1^ Dystonia can be due to inherited, acquired, or idiopathic causes. It contributes to impaired motor function, such as difficulties writing, speaking, or walking, and is more functionally disabling than spasticity. Moreover, it is often painful and can limit quality of life. While there are medical and surgical treatments for dystonia,^2-4^ existing interventions are often inadequate. Neuromodulation with deep brain stimulation (DBS) is well-established for certain types of dystonia, yet it is utilized in a very small percentage of the overall population affected by dystonia. There is emerging evidence that transcranial magnetic stimulation (TMS) may help manage dystonia symptoms, but studies to date have been mixed or shown only partial improvement, and this treatment is still considered investigational.^5^ Although dystonia has been studied for over a century, there remain large gaps in understanding its pathophysiology and neuroanatomical substrates. This limits the ability both to predict the development of dystonia and to identify effective treatments.

While lesion-induced dystonia is often associated with injury or dysfunction to the basal ganglia, lesions isolated to the cortex, cerebellum, and white matter can also result in dystonia, leading to the current belief that dystonia is a disorder of motor networks.^6, 7^ An evolving understanding of lesion-induced dystonia pathophysiology suggests that brain injury results in maladaptive plasticity, leading to abnormal neural reorganization of the cortico-basal-cerebellar network. This results in reduced selectivity for a desired motor pattern and decreased inhibition of other motor pattern generators, physically manifesting as the abnormal posturing and overflow movements characteristic of dystonia.^6, 8, 9^ Neuroimaging studies of dystonia in adults support this pathophysiologic model, by implicating differences of functional brain networks that span the basal ganglia, cerebellum, premotor cortex, sensorimotor, and frontoparietal regions.^10,11^

A recent study evaluating lesion-induced dystonia demonstrated most brain lesions in patients with dystonia involve the thalamus, basal ganglia, and cerebellum, further supporting a shared dystonia lesion network.^12^ Although there are few studies of brain networks in pediatric lesion-induced dystonia, a study investigating whole-brain structural networks in dyskinetic cerebral palsy, often associated with dystonia, found impaired network metrics of white matter integrity involving the sensorimotor, visual, and primary auditory areas.^13^

Elucidating the anatomy of dystonia lesion networks may inform clinical practice in the management of dystonia. For example, symptoms of acquired dystonia may not be evident at the onset of a brain insult, so identifying individuals at risk could lead to earlier initiation of treatments. Further, a better understanding of the dystonia-associated network could lead to the identification of more effective anatomical targets for therapeutic neuromodulation.^14^ However, understanding the neuroanatomy of idiopathic (non-lesional) dystonia is challenging as it is unclear if anatomical findings are functionally contributing to symptoms or represent mere correlations or downstream effects of having dystonia. Studying the anatomy of focal acquired brain lesions of individuals that subsequently develop dystonia has the potential to inform the causal underpinnings of dystonia.

Lesion network mapping is an approach that combines traditional lesion mapping of symptoms with structural and/or functional connectivity information derived from large normative ‘connectome’ datasets to evaluate lesion associated networks.^15^ These approaches have been applied to other lesional disorders to identify brain networks associated with clinical outcomes in tuberous sclerosis complex,^16^ cerebellar tumors,^17^ and stroke,^18^ among others, but has not been attempted in pediatric dystonia. Here, we utilize lesion network mapping in three separate pediatric patient cohorts with focal, acquired brain lesions who underwent standardized clinical assessments of dystonia more than 20 months after the lesion onset, along with a comparison group with early-onset acquired brain lesions without dystonia. Our goals were to evaluate both the lesion location associated with dystonia across different etiologies and the lesion associated networks of dystonia.

## Methods

### Patients

Four patient groups were included in this study – three had dystonia and one was a lesion comparison group. Etiologies contributing to dystonia included neonatal hypoxic ischemic encephalopathy (HIE), kernicterus, and stroke. All subjects were identified retrospectively from clinical records or from existing databases across three participating sites: Children’s Mercy Hospital, Kansas City, The Hospital for Sick Children (SickKids), Toronto, and the University of Iowa.

HIE. Patients in the HIE group were retrospectively identified through searching MRI brain reports of children at the Children’s Mercy Hospital main campus from 2010-2020.

Inclusion criteria included patients that were two years or older, had neonatal HIE, and had focal lesions evident on a clinically acquired brain MRI. These individuals were identified through review of electronic medical record brain MRI reports using the search terms: “hypoxic ischemic injury” OR “hypoxic ischemic encephalopathy” AND “putamen” OR “deep gray matter”. As the exact phrasing varied in some reports, an additional filter was added to include impressions within three words of “hypoxic ischemic injury” or “hypoxic ischemic encephalopathy,” which resulted in inclusion of similar phrases such as “hypoxic ischemic event” or “hypoxic-ischemic.” Chart review then identified which patients had a neonatal history of hypoxic ischemic brain injury and a current diagnosis of dystonia. Gross Motor Function Classification Scale (GMFCS) was recorded from chart data. Patients with other co-morbidities (e.g. sickle cell disease, Kearns-Sayre syndrome, or hypoxic event outside of the newborn period) were not included. Patients with non-focal lesions (global injury or atrophy) were excluded.

Kernicterus. The kernicterus patients were also identified retrospectively through searching patients seen at the Children’s Mercy Hospital Kernicterus Clinic from 2011-2020 who were two years of age or older and had a brain MRI available for review. The inclusion and exclusion criteria and methods for data extraction were otherwise similar to that of HIE patients.

Stroke. Patients with a history of arterial ischemic stroke diagnosed from birth to 18 years of age were retrospectively identified from the Toronto site of the Canadian Pediatric Ischemic Stroke Registry (CPISR).^19^ Inclusion criteria included available brain MRI imaging, a single infarct, and a clinical diagnosis of dystonia.

Comparison Patients. Comparison patients were derived from each site and the inclusion criteria included a focal brain lesion acquired before age 25 with no documented symptoms of dystonia. Their inclusion allowed us to evaluate whether regional lesion findings in the dystonia group were unique to dystonia patients versus occurred at sites that are commonly lesioned. Etiology for brain lesions in the comparison group varied but included primarily ischemic or hemorrhagic infarcts.

### Imaging and Lesion Analyses

Structural neuroimaging with MRI was obtained for each participant. The boundaries of the lesions were manually segmented for all scans using standard procedures.^20^ Lesions were manually traced onto the patient’s T1 native scan in FSL and then subsequently transformed into MNI152 space using ANTs.^21^ The anatomical accuracy of the native trace and the transformed lesion mask were confirmed and edited as needed by a neurologist (A.D.B. & R.G.M.).

Lesion-symptom mapping was used to identify regions of damage associated with dystonia relative to the comparison group. LESYMAP, a multivariate lesion-symptom mapping technique that uses sparse canonical correlation analysis for neuroimaging (SCCAN) was employed.^22^ We performed lesion-symptom mapping for all subjects together as well as separately by etiology.

Functional lesion network mapping was performed using each individual lesion volume as a ‘seed’ region in a normative resting state functional connectivity MRI (rs-fcMRI) dataset to generate a lesion-derived network. A pediatric normative rs-fcMRI dataset (ABCD1000) was used that consisted of 9-year-old typically developing participants (N=1000).^23, 24^ Rs-fcMRI data were processed in accordance with previously described methods.^25-27^

Each individual lesion ‘seeded’ a network map that showed the strength of connectivity from the lesioned site to the rest of the brain. Connectivity strength was represented as Fisher r-to-z scores to quantitatively demonstrate regions with positive and negative correlations with the average BOLD signal time course of the lesion volume. Brain regions with higher connectivity in relation to dystonia status were identified using a two-tail voxel-wise general linear model with 10,000 permutations and voxel-wise family-wise error (FWE) correction. This was implemented using FSL PALM, as previously described.^28, 29^

Structural lesion network mapping was performed using BCBtoolkit “*Disconnectome*” function.^30^ Similarly to the rs-fcMRI analysis described above, each lesion was used to seed tractography streamline maps from a normative population of 100 Human Connectome Project (HCP) participants. Streamlines intersecting each voxel were derived from pre-defined tractograms, converted into binary masks and then averaged across all participants. As described above for the fcMRI analysis, all lesion-seeded voxel-wise streamline maps and corresponding dystonia status were included in a permutation-based voxel-wise two-tail linear model via FSL PALM with FWE correction of multiple comparisons, as performed with functional network data.

## Results

31 individuals with acquired dystonia and 93 comparison subjects were identified. The initial search of patients with HIE identified 408 MRI brain reports, of which 10 patients were identified that met inclusion criteria for neonatal HIE with a discrete brain lesion and a current diagnosis of dystonia. Chart review of kernicterus patients identified 37 patients, of which 11 had a brain MRI and a confirmed diagnosis of dystonia. Three patients were excluded due to poor image quality, leaving a total of eight patients with kernicterus for inclusion in the study. A query of the Canadian Pediatric Ischemic Stroke Registry identified 74 patients with dystonia, 53 of which were excluded for having multiple lesions. Eight patients were excluded due to poor imaging quality, leaving 13 patients with dystonia and a single ischemic stroke for inclusion in the study (Figure 1). Brain injury occurred in the perinatal and neonatal period for the HIE and kernicterus groups, respectively, and from birth to 14.83 years (mean 4.86 years) in the stroke group (Table 1). All patients with kernicterus and HIE included in the study had a dystonia-predominant cerebral palsy phenotype, with little to no documented spasticity. Mean Gross Motor Functional Classification Scale (GMFCS) in the HIE group was 4 (range 2-5) and 4.38 in the Kernicterus group (range 1-5). Patients in the stroke group had a documented mix of dystonia and spasticity, measured by the Hypertonia Assessment Tool (HAT) (mean 3, range 1-6) and the Pediatric Stroke Outcomes Measure (PSOM) (mean 2, range 0.5-3). A total of 93 comparison patients with early-onset focal acquired brain lesions were included in the study (age range at time of injury birth-24.97 years, mean 12.86 years) (Table 1).

**Table 1:**
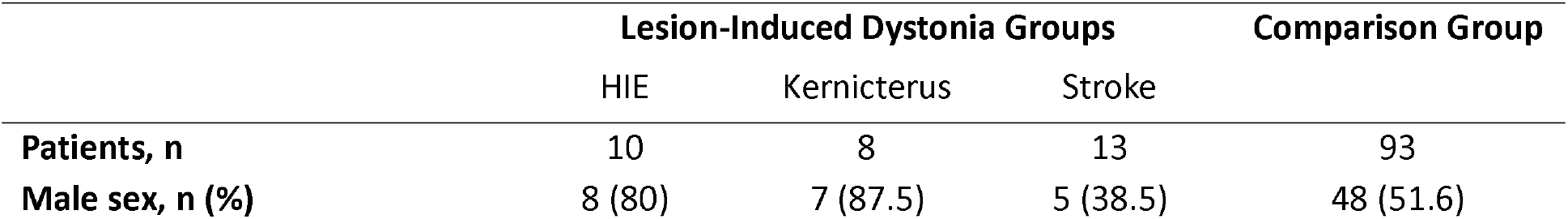
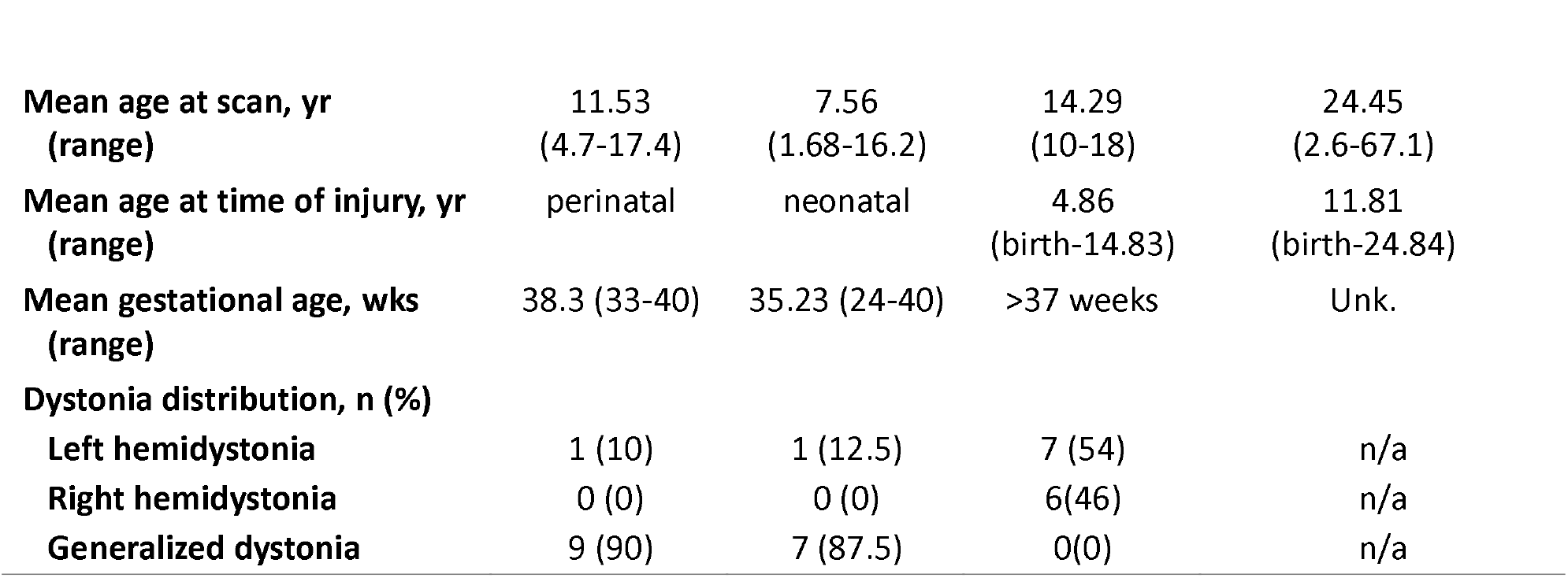
Demographic Data

|  | Lesion-Induced Dystonia Groups |  |  | Comparison Group |
| --- | --- | --- | --- | --- |
|  | HIE | Kernicterus | Stroke |  |
| **Patients, n** | 10 | 8 | 13 | 93 |
| **Male sex, n (%)** | 8 (80) | 7 (87.5) | 5 (38.5) | 48 (51.6) |

Table 1: Demographic Data
|  |  |  |  |  |
| --- | --- | --- | --- | --- |
| **Mean age at scan, yr**<br>**(range)** | 11.53<br>(4.7-17.4) | 7.56<br>(1.68-16.2) | 14.29<br>(10-18) | 24.45<br>(2.6-67.1) |
| **Mean age at time of injury, yr**<br>**(range)** | perinatal | neonatal | 4.86<br>(birth-14.83) | 11.81<br>(birth-24.84) |
| **Mean gestational age, wks**<br>**(range)** | 38.3 (33-40) | 35.23 (24-40) | >37 weeks | Unk. |
| **Dystonia distribution, n (%)** |  |  |  |  |
| **Left hemidystonia** | 1 (10) | 1 (12.5) | 7 (54) | n/a |
| **Right hemidystonia** | 0 (0) | 0 (0) | 6(46) | n/a |
| **Generalized dystonia** | 9 (90) | 7 (87.5) | 0(0) | n/a |

**Figure 1.**
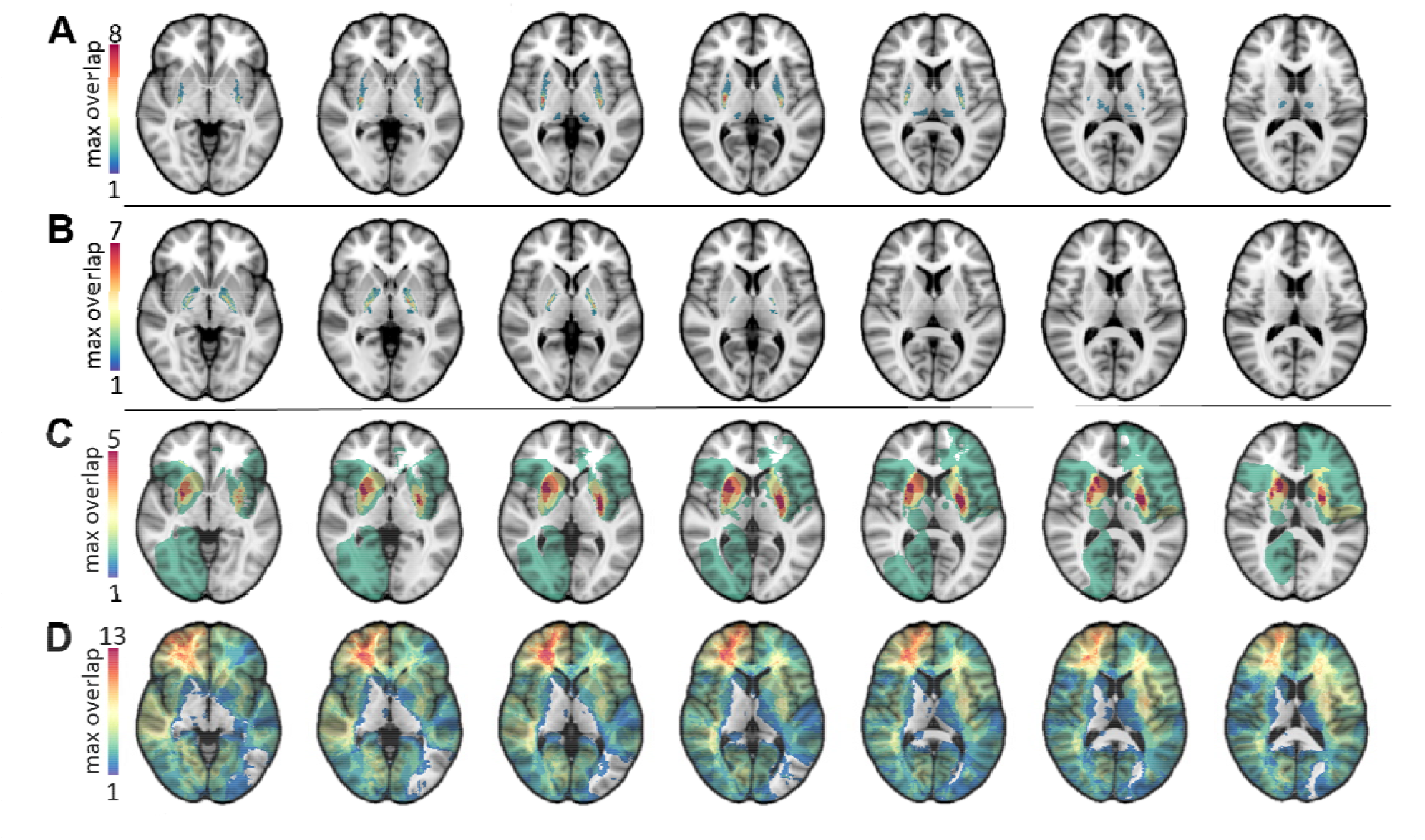
A-D. Lesion masks of all participants in this study are shown, separated by group. The lesion masks of the 10 participants with HIE are shown in 1A; the region of peak overlap in the putamen contained 8 lesions. The overlap of kernicterus participants is shown in 1B. The maximum number of overlapping lesions (7) was located in the globus pallidus. The peak lesion overlap of the stroke participants was in the putamen (5), shown in 1C. The comparison group had a peak lesion overlap in the right frontal lobe (max overlap=13, MNI 30 41 0) (1D).

### Lesion mapping results

Lesion overlap maps for the HIE, kernicterus, stroke and comparison groups are shown in Figure 1A-1D. All lesions in the HIE group were located in the putamen (peak overlap N = 8 of 10, MNI 29, -14, 5), and lesions in the kernicterus group were located primarily in the globus pallidus (N = 7 of 8, -18, -8, -1). Lesions in the stroke participants with dystonia were more heterogeneous, involving the right or left cortical and subcortical gray and white matter. The peak site of lesion overlap was in the putamen bilaterally (peak overlap N=5 of 13, MNI 25, 9, 5 & -28, -6, 5 and caudate MNI 18, 11, 15). The lesion overlap for comparison subjects covered several brain areas, including cortical and subcortical gray matter with the highest overlap in the right frontal lobe (N = 13 of 93, MNI 30, 41, 0).

Lesion-symptom mapping with all subjects was significant in showing an association of lesions of the putamen as having the highest association with dystonia (r = 0.423, p< 0.001) (Figure 2A). When performed separately by etiology, each analysis was also significant (HIE: r = 0.64, p <0.001, peak MNI = 29, -10, 3; kernicterus: r = 0.61, p <0.01; peak MNI = 23, -9, 0; stroke: r = 0.50, p <0.01; -25, 0, 6) (Figure 2).

**Figure 2.**
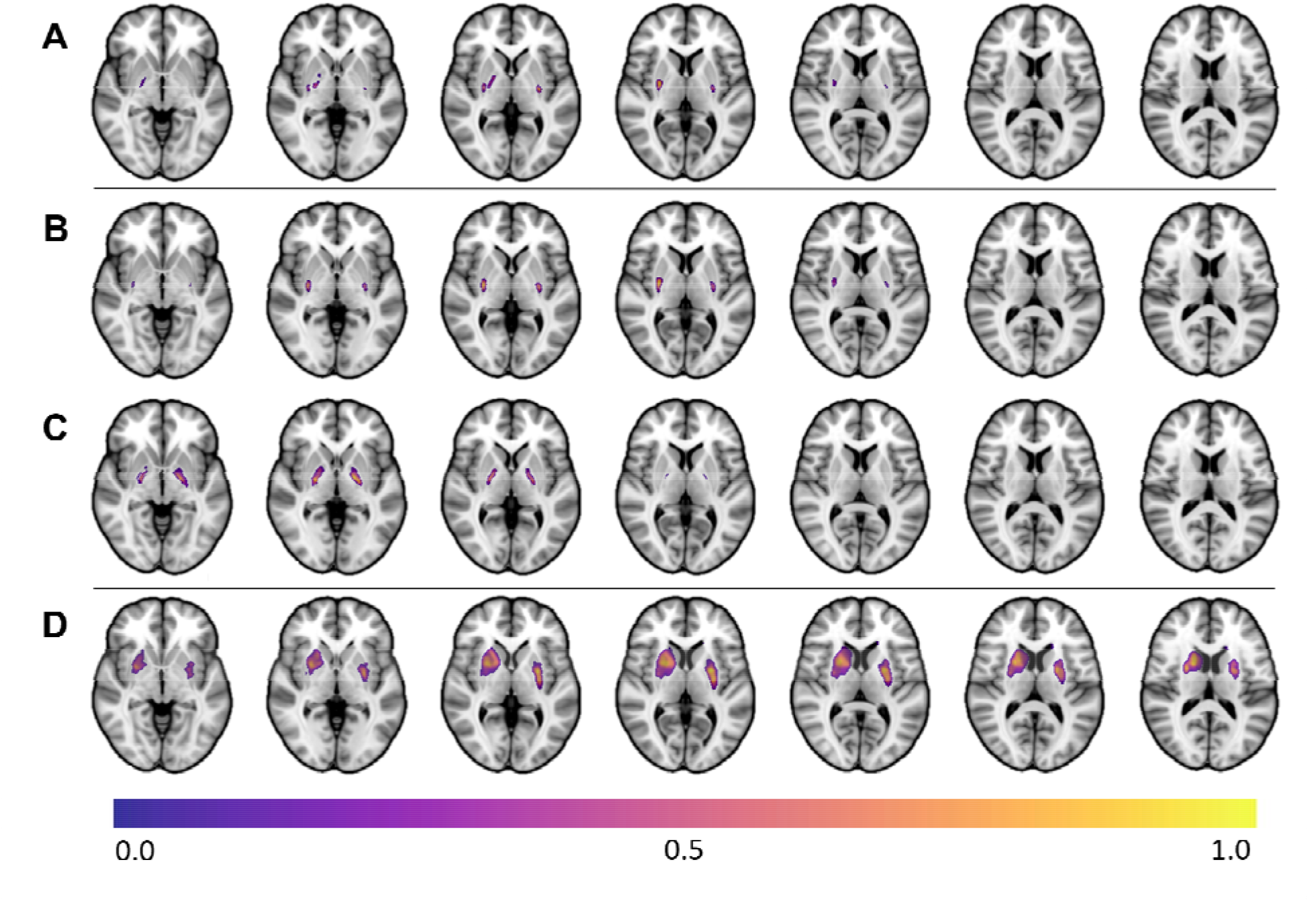
A-D. The lesion-symptom mapping analysis was first conducted with the entire sample (2A, r = 0.42, p < 0.001) and showed regions of peak association with dystonia in the putamen and globus pallidus. Lesion symptom maps for each etiology analyzed independently are also shown, including hypoxic ischemic encephalopathy (2B, r = 0.64, p < 0.001), kernicterus (2C, r = 0.61, p < 0.001), and stroke (2D, r = 0.5, p < 0.001). Voxels are scaled to values between 0 and 1, with greater values indicating a greater association with dystonia.

Combining all dystonia groups (kernicterus, HIE, and stroke) further defined a shared dystonia lesion network map involving the globus pallidus, posterior putamen, and thalamus (p = 0.0001; peak voxel 24, -10, 0), as well as the primary motor cortex (46, -10, 42), cerebellum (0, -54, -24; 10, -54, -26), and brainstem (8, -24, -16) (Figure 3). When displayed on the cortical surface, the most robust network connection to dystonia lesions is in the primary motor cortex (M1), dorsal anterior cingulate, and insula. The peak regional findings within the motor cortex correspond to the inter-effector nodes of the somato-cognitive action network (SCAN) (Figure 3).^31^ To evaluate the lesion-derived dystonia network similarities to SCAN further we compared the spatial correlation of the dystonia network to a network map seeded by effector versus inter-effector nodes of the motor cortex. The dystonia network was more similar to the SCAN network (r = 0.56 versus 0.38, p < 0.001). In addition to SCAN, the other fcMRI network with a similar distribution to the dystonia network was the cingulo-opercular/action-mode network (CON/AMN) (r = 0.57). The spatial correlation with SCAN AND CON/AMN was significantly higher than with any of the canonical networks in the Yeo 7 network parcellation, with ventral attention and somatomotor networks having the highest spatial correlation to our results (r = 0.40 & 0.23, respectively). These results are provided in Figure 1 of supplemental material).

**Figure 3.**
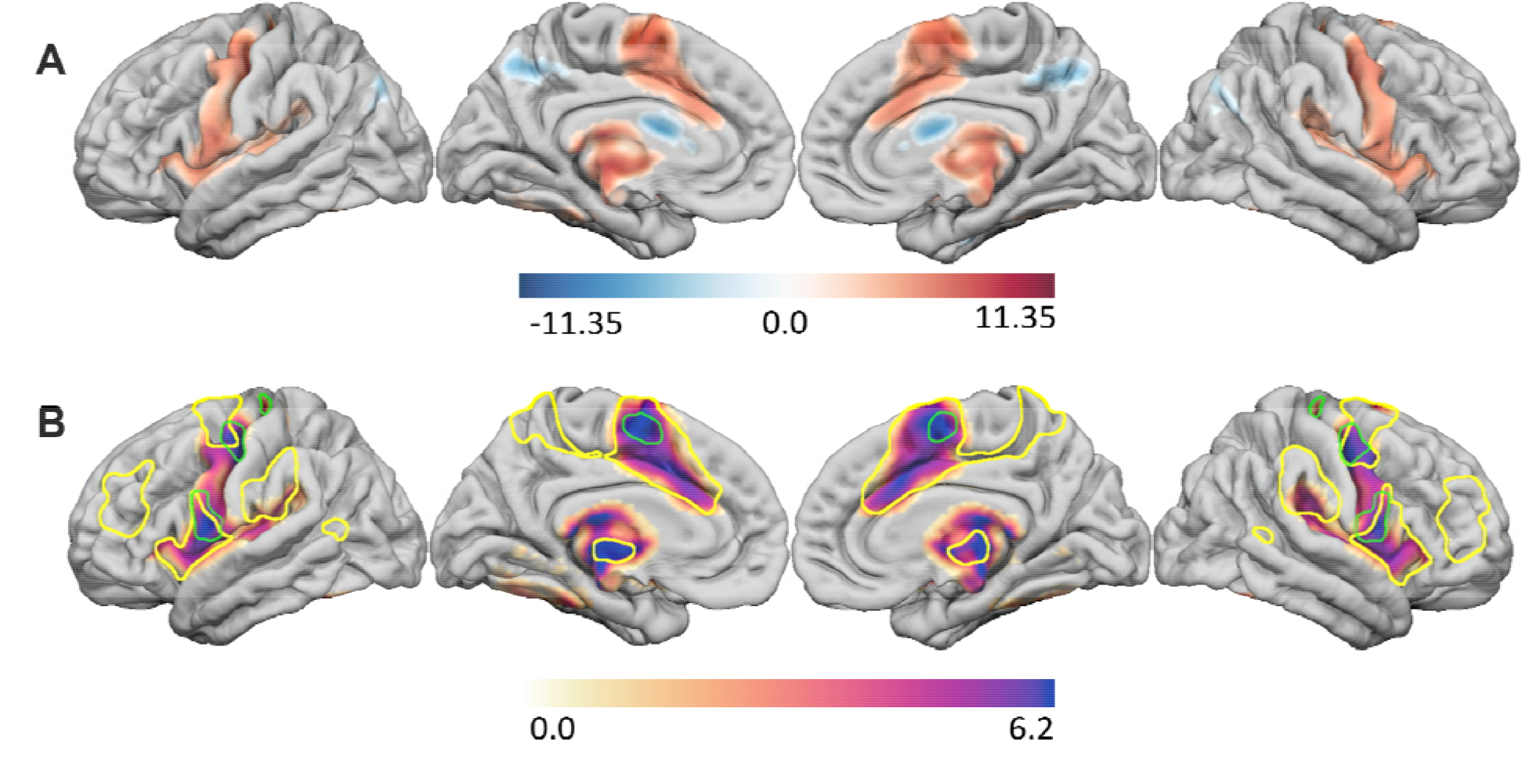
Dystonia lesion network maps. A) The functional dystonia lesion network map for all etiologies combined is shown in red (scaled between 0 – 11.35) and the structural dystonia lesion network map is shown in blue (scaled between –11.35 and 0.0). Only regions reaching statistical significance after correction for multiple comparisons are shown. The voxel range is T = -9.4 – 12.33, p = 0.0001 for grey matter and T = 0 – 8.65, p = 0.0029 for white matter. B) The functional lesion network map has regional peaks that appear to correspond with the inter-effector nodes of the somato-cognitive action network (green outlined areas) and cingulo-opercular/action-mode network (yellow outlined areas).

Analysis of white matter regions significantly more connected with dystonia lesions relative to comparison lesions revealed significantly greater connectivity between dystonia lesions and the superior primary motor and primary sensory cortex, as well as the brainstem and cerebellum. The structural lesion network map findings reaching significance overlapped most prominently with the corticospinal and parietopontine tracts, often at sites of overlap between these tracts.^32^ The peak voxel in this structural lesion network map was 20, -6, 0; p = 0.0029) (Figure 3).

Lesion network mapping results were also generated independently for each lesion etiology and substantially overlapped with the combined results. The functional lesion network maps derived from each individual etiology also reached statistical significance (each p < 0.001 with FWE correction) and were highly correlated with one another, in the range of r = 0.76 – 0.95 (Figure 2, supplemental material). The functional lesion network maps for all lesion etiologies without FWE correction are also provided as supplemental material to demonstrate the spatial similarity (Figure 3, supplemental material). The structural lesion network maps derived from each etiology independently were also generated and two of three reached significance (kernicterus p = 0.003; HIE p = 0.03; stroke p = 0.067). The two structural lesion network maps that reached statistical significance were spatially similar to the combined result, with spatial correlations of 0.84, 0.76 for kernicterus, HIE, respectively.

## Discussion

This multicenter study evaluated lesion location and structural and functional network connectivity in three distinct etiologies of lesion-induced dystonia. We showed that damage to the putamen and globus pallidus were associated with dystonia, in line with prior findings.^12^ Next, we identified a dystonia lesion network that was common across the different lesion etiologies. A brain-wide functional network involves the primary motor cortex, lentiform nucleus, thalamus, and cerebellum. Interestingly, the functional network derived based on connectivity to dystonia lesions includes the recently described somato-cognitive action network (SCAN), a network that is located primarily within the primary motor cortex (M1) in regions less directly tied to movements, but thought to have more of an integrative role in movement.^31^ Regions of the cingulo-opercular / action-mode network (CON/AMN) were also included. This network is implicated in motor planning and is sensitive to feedback from actions.^33^ These network findings were further reinforced with structural network data, which implicates disrupted connectivity of white matter along the intersection of the corticospinal and parietopontine tracts, which could serve to disrupt somatomotor connectivity with the brainstem and cerebellum.

The importance of these results is multifold. First, this is the first association between dystonia and the SCAN, to our knowledge. The SCAN contains inter-effector regions which intercalate in the traditional M1 motor homunculus. The inter-effector regions of the SCAN are hypothesized to play a role in higher order coordination of movements as opposed to the effector sites in M1, which represent the classic somatotopic homunculus. The inter-effector regions are highly connected with each other and also connect with the action-mode network (AMN),recently renamed from the cingulo-opercular network (CON),^33^ which includes the dorsal anterior cingulate and anterior insula, and is responsible for activating performance of specific tasks and error correction.^34^ Our dystonia lesion network demonstrates regional peaks of functional connectivity to M1, corresponding to the identified inter-effector nodes in the SCAN, and also peaks in the anterior cingulate and insula, corresponding to the CON/AMN. The involvement of these networks in the dystonia lesion network is not surprising. Dystonia is typically activated by attempts at voluntary movements, a task which would logically require both the activation of the SCAN for higher order movement integration and the CON/AMN for task activation and error correction. In this way, the dystonia lesion network revealed by this study gives insight into the functional underpinnings of dystonia.

In addition to the association with known functional networks, our structural and functional connectivity data support the importance of the cortico-basal ganglia-cerebellar network in dystonia. These structures are proposed to underlie the development of motor and cognitive functions, including dystonia.^35^ While the basal ganglia have long been implicated in dystonia, there is increasing awareness of the role of the cerebellum.^36, 37^ Similarly, alterations of sensorimotor cortex connectivity are often implicated in dystonia. Although rarely seen in childhood, studies of focal dystonia in adults have demonstrated alterations in connectivity to the motor cortex in spasmodic dysphonia,^38^ cervical dystonia,^39^ and writer’s cramp.^40, 41^ EEG connectivity studies have found abnormalities in the cortical electrophysiological patterns in patients with dystonia,^42^ including patients with dystonic cerebral palsy.^43^ Our study highlights the importance of the cortico-basal ganglia-cerebellar network in lesion-induced dystonia, through both functional and structural network involvement of these regions.

Finally, knowledge of the dystonia lesion network gained from this study could have important implications for advancing dystonia diagnosis and treatment. Children with acquired brain lesions connected to the dystonia lesion network may be at increased risk of developing dystonia, guiding increased surveillance for dystonia to facilitate earlier diagnosis and intervention. Brain regions within the dystonia lesion network could also serve as novel sites for deep brain stimulation (DBS) or non-invasive brain stimulation. The target for DBS in dystonia is typically the globus pallidus internus (GPi).^44^ However, in patients with lesion-induced dystonia, their GPi is frequently injured or atrophied, and may not be the ideal site for DBS lead implantation. Others have speculated that cerebellar stimulation may provide an alternative site for DBS in dystonia patients.^45^ Our results support the possibility of cerebellar DBS as an anatomical target capable of modulating a dystonia lesion network by demonstrating strong connectivity between the deep gray matter and cerebellum in all patient groups. Similarly, children with lesion-induced dystonia may benefit from stimulation to cortical regions within the dystonia lesion network, potentially providing an option for non-invasive brain stimulation in dystonia treatment.^46^

Interestingly, these network associations with dystonia may differ depending on the type of dystonia. A recent cervical dystonia lesion network mapping study identified that lesions with connectivity to the cerebellum and the somatosensory cortex were associated with dystonia.^47^ While our study also found cerebellar connectivity in lesions associated with dystonia, the connections were to the anterior vermis, which is different than what was reported previously.

Further study will be needed to determine the extent of variability across different lesion-induced types of dystonia.

There are some limitations to this study. We cannot rule out the possibility that some individuals in the comparison group could have had undocumented symptoms of dystonia, though we could not find any records of dystonia and would expect that this cohort should not have been enriched in dystonia in the way our dystonia groups were. Lesion symptom mapping in all the patients in this study found lesions in the deep gray matter were associated with dystonia (putamen for HIE group, putamen and caudate for stroke group, and globus pallidus for kernicterus group). Many patients with isolated periventricular or cortical based injuries can also develop dystonia^48^ and these patients were not well represented in this study. As the primary motor cortex and associated white matter tracts connecting this region to the deep gray matter are part of the dystonia lesion network, our results raise the hypothesis that patients with dystonia and isolated lesions in the cortex or white matter involve portions of this network.

Future studies including a larger patient population with more heterogenous lesion locations could explore this possibility. Additionally, these studies may reveal dystonia lesion network subtypes predictive of specific dystonia phenotype and response to a particular medication or surgical intervention, such as DBS.

## Conclusion

In summary, results from this study demonstrated that damage to the putamen and globus pallidus were associated with dystonia, but further found that lesions associated with dystonia are connected to a common structural and functional brain network including the motor cortex, dorsal anterior cingulate, deep gray matter, and cerebellum. This dystonia lesion network overlaps with the SCAN and CON/AMN, which are involved in action selection and integrating motor activity with other modalities of information. Results from this study enhance our understanding of dystonia pathophysiology and could be useful to aid in dystonia surveillance and identification of novel sites for deep brain stimulation. Future expanded studies including more patients with lesion-induced dystonia will refine the dystonia lesion network and identify network subtypes to further guide dystonia treatment.

## Data Availability

All data produced in the present study are available upon reasonable request to the authors.

## Acknowledgements

This work was supported by NIH NICHD T32HD069038 (RGM), The Auxilium Foundation (ND), NIMH K23MH120510 and the Simons Foundation Autism Research Initiative (AC), NIH NICHD P50 HD103525 (CDS), and NIH NINDS 3R01NS114405 (AB).

## Author Contributions

RGM, ND, CDS, and AB conceived of the project idea and designed the study. RGM, ND, AR, JB, and AC acquired and analyzed the data. RGM, ND, and AB drafted a significant portion of the manuscript and figures. All authors contributed to manuscript revisions.

## Conflicts of Interest

No authors have any conflicts of interest to report.

## Notes

### Competing Interest Statement

The authors have declared no competing interest.

### Author Declarations

IRB of Children's Mercy Kansas City gave ethical approval for this work.

