## Supplemental Figures for "Network localization of pediatric lesion-induced dystonia"

Supplemental Figure 1: Correlation matrix between functional dystonia lesion network (dystonia-GM) and other establish brain networks, including the inter-effector nodes of the SCAN network (IEN), the effector nodes of the motor cortex (HandFaceFoot), and the action-mode network/cingulo-opercular network (AMN-CON).  We also included other Yeo 7 networks (VIS, SMN, DAN, VAN, LIM, FPN, and DMN).  Correlation r values are displayed in in a color gradient from 0 – 1, with darker colors indicating a greater r value. Correlation with the functional dystonia lesion network was highest with the inter-effector nodes of the SCAN network (r = 0.56) and the AMN-CON (r = 0.57).
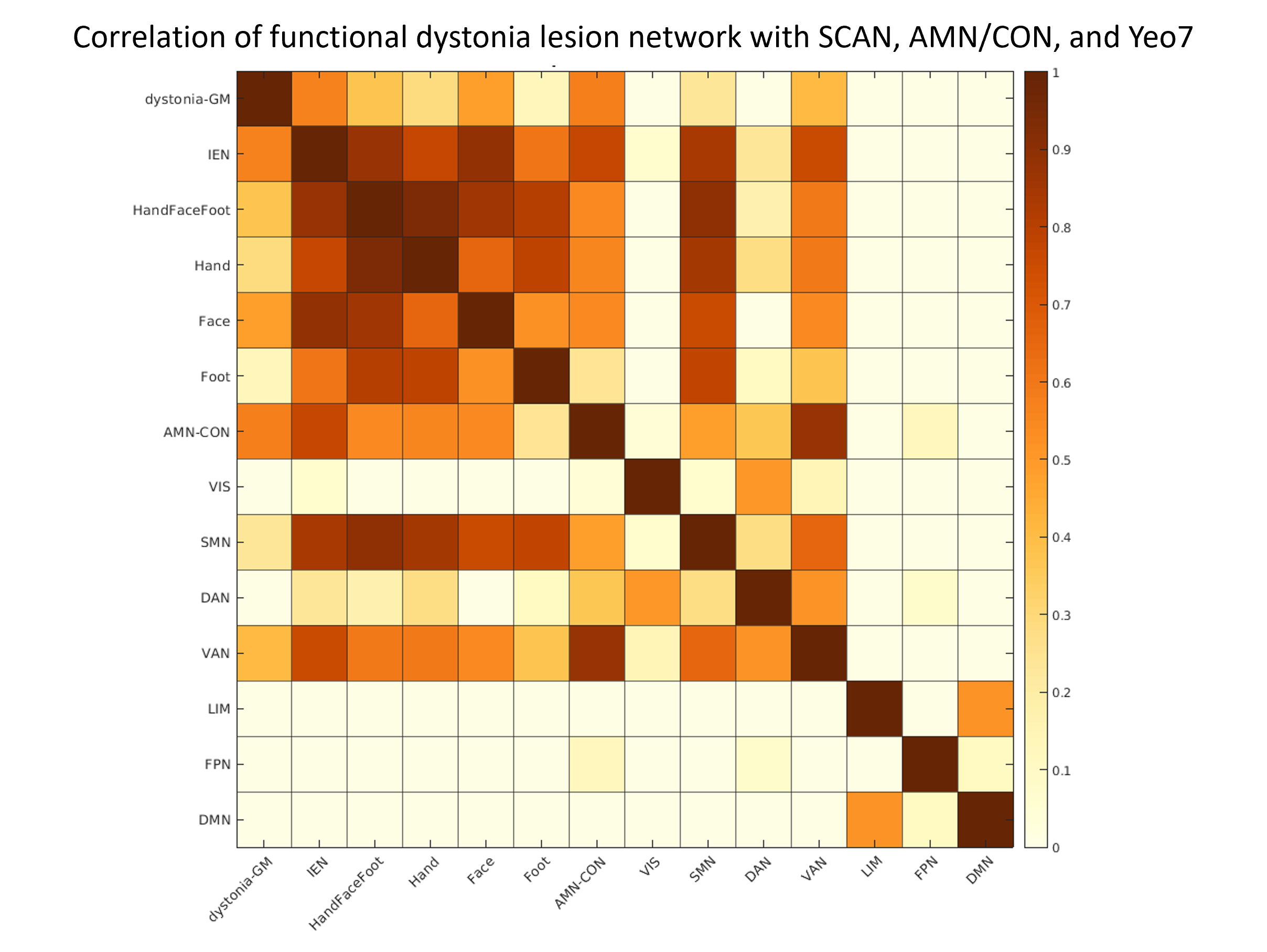

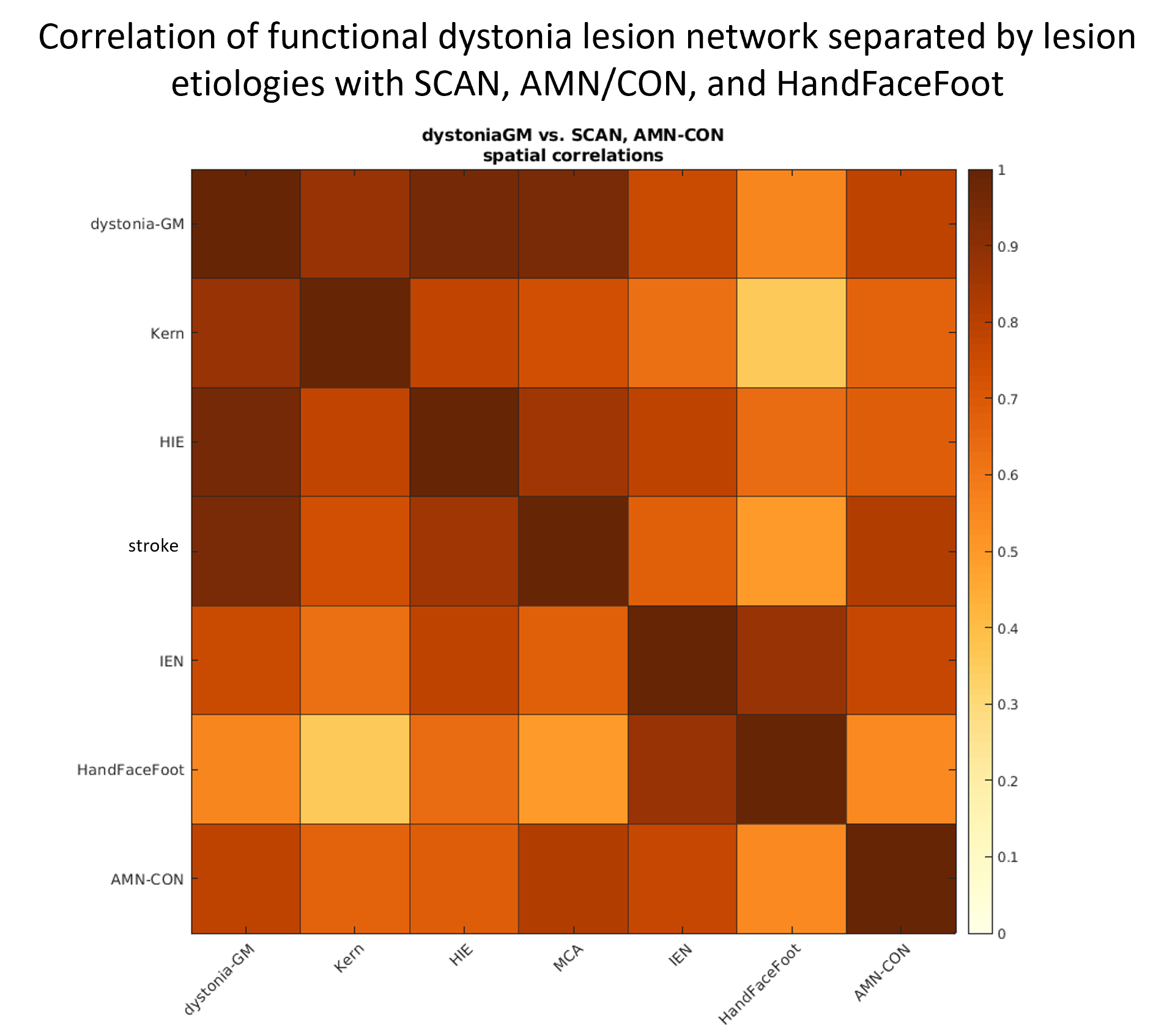

Supplemental Figure 2: Correlation matrix between functional dystonia lesion network (dystonia-GM), functional dystonia lesion network divided by lesion etiology, inter-effector nodes of SCAN network (IEN), the effector nodes of the motor cortex (HandFaceFoot), and the action-mode network/cingulo-opercular network (AMN-CON). Correlation r values are displayed in in a color gradient from 0 – 1, with darker colors indicating a greater r value. Functional lesion networks were highly correlated between lesion etiologies (r = 0.76 – 0.95) and also highly correlated with the inter-effector nodes of SCAN and the AMN-CON.  Kern = Kernicterus, HIE = hypoxic ischemic encephalopathy

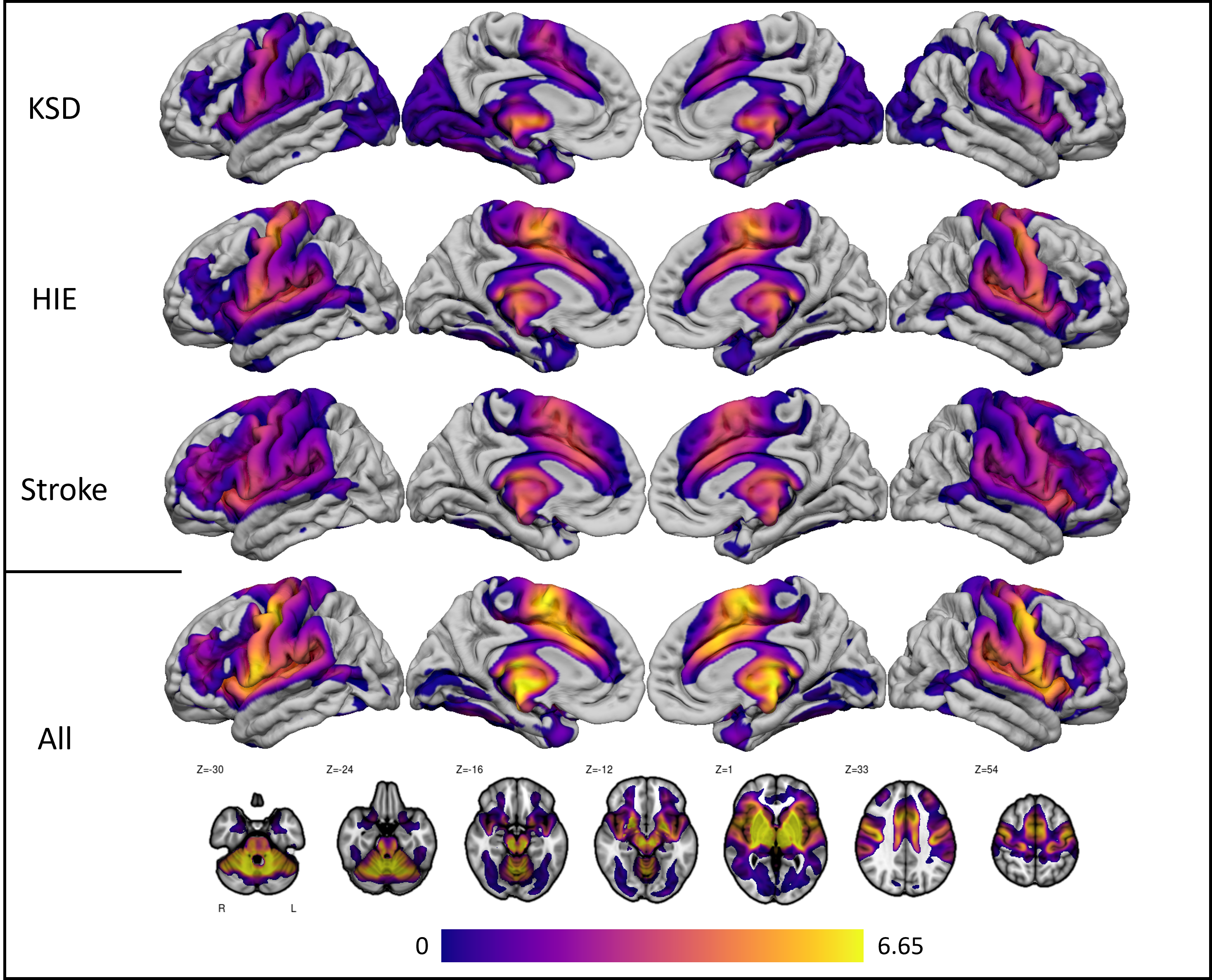

Supplemental Figure 3: Functional lesion network mapping results separated by individual lesion etiology.  Most results in individual lesion etiology lesion network maps did not survive correction for multiple comparison and are therefore shown uncorrected for multiple comparisons to demonstrate similarities in lesion network maps between lesion etiologies.
